# DETECTION OF AUTOLOGOUS BLOOD DOPING

**DOI:** 10.1101/2020.07.04.20146217

**Authors:** Christer Malm, Irene Granlund, Michael Hall, Pernilla Lindén, Magnus Ericsson, James I. Langridge, Lee A. Gethings, Chris Hughes, Nelson Khoo

## Abstract

**Objectives:** Autologous blood transfusion (ABT) enhances athlete’s performance, is banned as doping by the World Anti-Doping Agency (WADA). Currently, there is no implemented detection method for ABT. Transfusion of one’s own, long-term cryopreserved red blood cells (cryo-RBC) immediately increases circulating RBC count, hemoglobin mass, blood volume and oxygen carrying capacity, resulting in enhanced physical performance. Functional viablity of cryo-RBC are maintained for decades, but storage lesions lead to removal of damaged RBC from circulation days after transfusion, with remaining circulating cryo-RBC displaying normal half-life.

**Methods:** The cytosolic RBC peptidome from 22 human subjects (12 men and 10 women) was analyzed by UHPLC-MS/MS before and after ABT with cryo-RBC. As a control group and for investigation of confounders, 14 elite athletes and 5 recreational subjects were sampled multiple times, also at high altitude.

**Results:** Here we report alteration in the cytosolic peptidome of circulating RBC weeks after ABT, discriminating doped from non-doped human subjects. A valid discriminating multivariate model (OPLS-DA) based on <200 peptides was accomplished (R^2^/Q^2^ = 0.88/0.59, P CV-ANOVA < 0.0001, ROC AUC = 0.97). Models did not show bias for sex, high altitude or elite endurance training and racing.

**Conclusion:** Identified peptides with low intra- and inter-individual variation, and high multivariate model weight and probability scores, create a direct method for the detection of autologous blood doping.

## Introduction

Autologous blood transfusion (ABT) enhances the athlete’s performance,^1^ and is viewed as doping, thus banned by the World Anti-Doping Agency (WADA); but with no implemented direct detection method,^2^ it remains difficult to survey by sporting authorities. Because cold-stored (+1° to +4°C) blood is viable for a maximum of 42 days,^3^ during which time the athlete’s physical performance is compromised from blood donation,^4, 5^ and erythropoietin is readily detected;^6^ the blood doping method of choice for cheating athletes is transfusion of one’s own, long-term cryopreserved red blood cells (cryo-RBC). Upon transfusion, increases are immediately observed in circulating RBC count, hemoglobin mass,^7^ blood volume and oxygen carrying capacity,^8^ resulting in enhanced physical performance.^1, 9, 10^ Cryo-RBC in storage maintain their function for more than a decade,^11, 12^ but display storage lesions altering physical properties^13, 14^ of both cytosolic^15^ and membrane ^16^ proteins, leading to an expedited removal of damaged cryo-RBC from circulation 6 h to 48 h after transfusion.^17, 18^ The remaining circulating cryo-RBC display normal half-life^19^ despite observable altered metabolic profiles^20^ and irreversible cytosolic protein alterations, derived from the process of freezing and thawing.^21^

Today, indirect detection of ABT exist using the hematological module of the Athletes Biological Passport (ABP). The ABP uses longitudinal monitoring of standard hematological parameters to identify possible blood manipulations,^22^ and can be considered less effective than anticipated in detecting ABT.^23, 24^ Also, the ABP is affected by exercise^25^ as well as increased fluid intake.^26^ Moreover, several tests claim to detect ABT via: immunological reactions,^27^ hematological variables,^28^ phthalate/metabolites^29^ and CO rebreathing^30, 31^, but no test is implemented for doping detection.

A direct and valid detection method for ABT remains elusive and unavailable,^1, 32, 33^ but must include intracellular biomarkers, as all cell-surface alterations, including storage-induced,^34^ initiate rapid removal of cryo-RBC from circulation by spleen macrophages,^18^ thus circumventing the effectiveness of such markers for detection of ABT.

Proteomic and bioinformatic methodologies allows detection of sample differences in minutiae across subjects and chronology. We proposed that screening the cytosolic fraction of circulating RBC (RBCc) sampled before and after cryo-RBC transfusion, will provide markers specific to ABT. Subsequently, applying liquid chromatography tandem mass spectrometry (LC-MS/MS) analysis will generate biomarkers (proteins and/or peptides), differentiating the two states of pre- and post-transfusion, defining Clean and Doped samples, respectively. In addition, identified biomarkers will be curated to remove those markers biased for sex, exercise, hypoxia and high-altitude exposure.

## Method (Appendix 10)

### Subjects

Autologous blood transfusion of eight men (age 29; 24-36 years [mean; min-max]) and seven women (age 27; 23-38 years) with peak oxygen consumption (VO_2peak_); Men (4.3; 3.2-5.0 L·min-^1^/53; 33-59 mL·min-^1^·kg-^1^) and women (3.0; 2.4-3.6 L·min-^1^/48; 45-56 mL·min-^1^·kg-^1^) with five recreational Control subjects sampled in parallel (Appendix 2, 3 & 9). Fourteen international level elite endurance athletes investigated confounders by resting blood samples at: Sea Level (<80 m), High Altitude Training (>1500 m), Hypoxic Tents (>10 h/day, 14 days, 3-4000 m) and in the Hypoxia group up to 28 days after a multisport race (700 km in 4.5 days reaching altitudes up to 4500 m, Appendix 2, 3 & 9).

### TRANSFUSION

Women donated one (450 mL) and men two (900 mL) units of whole blood as s described in detail previously ^1, 18^. Whole blood was collected and glycerolized RBC stored at -80°C until transfusion.

### SAMPLING

In total, participants (21 men and 15 women, Appendix 2, 3 & 9) gave 307 venous blood samples, where 85 were in a Doped state after cryo-RBC transfusion, 173 in a Clean state and 49 samples taken directly from the Blood Donation Bag (timeline for Transfusion indicated in Figure 1). Some samples were used multiple times (noted as Batch I, II and III in Tables and Appendices), resulting in 392 LC-MS/MS sample injections (Appendix 2, 3 & 9).

**FIGURE 1.**
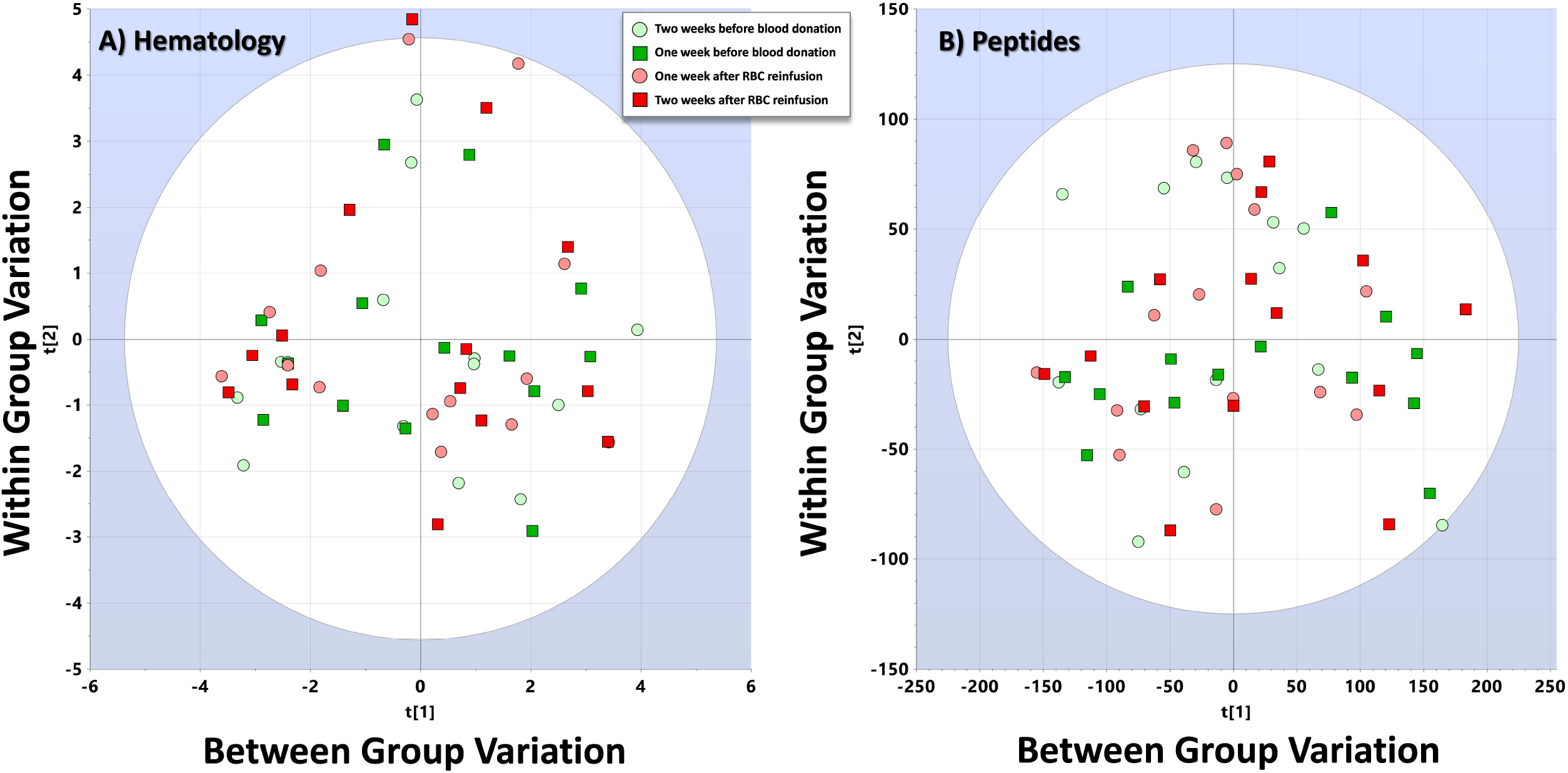
Hematology

### Preparation

Erythrocytes were lysed, the cytosol isolated and hemoglobin removed before determination of protein concentration. Pre-digest internal peptide standard was added before trypsin digestion. Clean-up of trypsin digest samples was performed by C18 and dried samples were stored at -80°C.

### Nano-UPLC-MS/MS Analysis

Internal retention time standards, pre-LC internal standard and TFA were added before injection using a nanoACQUITY UltraPerformance UPLC™ (Waters). Separated peptides were analyzed using a Synapt G2™ or G2Si™ (Waters). Initial data evaluation used the MassLynx™ software (Waters).

### Data processing and analysis

Raw data processing, alignment, peptide identification, quantification (relative abundance) and comparative analysis were performed using Progenesis QI for Proteomics (Waters), Protein Lynx Global Server (Waters) and the open-source Skyline (https://skyline.ms). For peptide identification a curated human proteome database (Swiss-Prot) was used, acquired from Uniprot (www.uniprot.org).

### Statistics

Mixed Model using Restricted Maximum Likelihood (REML) and multivariate analysis (MVA),^1^ with Principal Component Analysis (PCA), Orthogonal Projections of Latent Structures (OPLS)^35^, and its discriminant analysis OPLS-DA^36^ were applied. Samples from blood donation (Clean) and after transfusion (Doped), High Altitude Training, Hypoxic Tents and Elite Endurance Athletes created MVA models (Appendix 5 & 9). Regression (R^2^) and prediction (Q^2^) were investigated, with valid R^2^ and Q^2^ set to > 0.60, ^35, 37, 38^ in three datasets: Training (Men), Testing (Women) and Validation (Men and Women) for Screening, Selection and Validation of Biomarkers (Appendix 5 & 9).

### Confounders

Data from two LC-MS/MS analyses which differed to the biomarker screening dataset (Appendix 6), were used to create four OPLS-DA models investigating confounders. Appendix 7: 1) If peptides separating fresh and cryo-stored RBCc could separate Clean from Doped subject 2) Bias for Elite Endurance training and 3) Summary model (four Y -variables) of Clean, Doped, Elite Athletes and High-Altitude Training. Appendix 8: Bias for High Altitude Training and Hypoxic Tents.

## Patient and Public Involvement Statement

Participants in the study were recruited locally. Each participant was given continuous feedback. Lectures informed staff, participants and colleagues of the study objectives.

## Ethical considerations

The Ethics Board for Northern Sweden approved the study (DNr: Ö1-2009, 08-196M, 2011-408-32M), and transfusions performed in a hospital clinical. Risks reviewed in Appendix 1. The study was conducted in accordance with the WMA Declaration of Helsinki 2013. Data is encrypted and stored on security protected servers.

## Results (Extensive models in Appendix 5 & 9)

Normal distribution of samples is visualized in a PCA model for Hematological (Figure 1A) and Proteomic (Figure 1B) data.

### Hematology

Autologous blood transfusion changed hematological variables (Figure 1, Table 3, Appendix 3 and 4), but with large individual variations, making regression analysis and prediction of samples to Clean (Before blood donation) or Doped (after transfusion of cryo-RBC) unreliable (Figure 2 and 3, Appendix 3 and 4). The visualized OPLS-DA analysis (Figure 3) indicate models with low R^2^ (<0.6) and Q^2^ (<0.2) in the Summary of Fit plot, and sensitivity and specificity displayed in the ROC curve, where AUC for Men = 0.97 and Women = 0.79. Selection for prediction by VIP Pred indicate different variables of importance for men and women.

**FIGURE 2.**
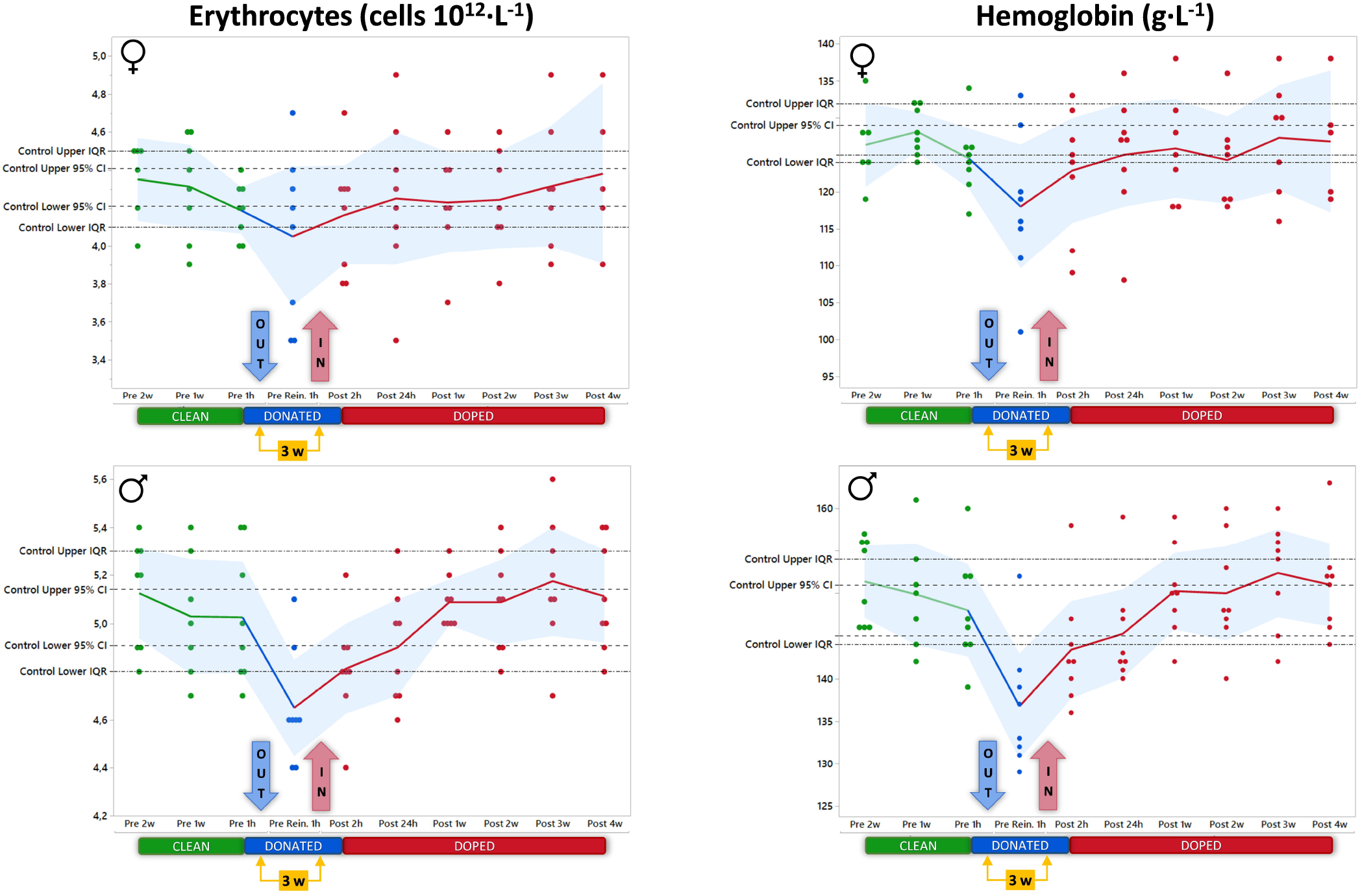
PCA models

**FIGURE 3.**
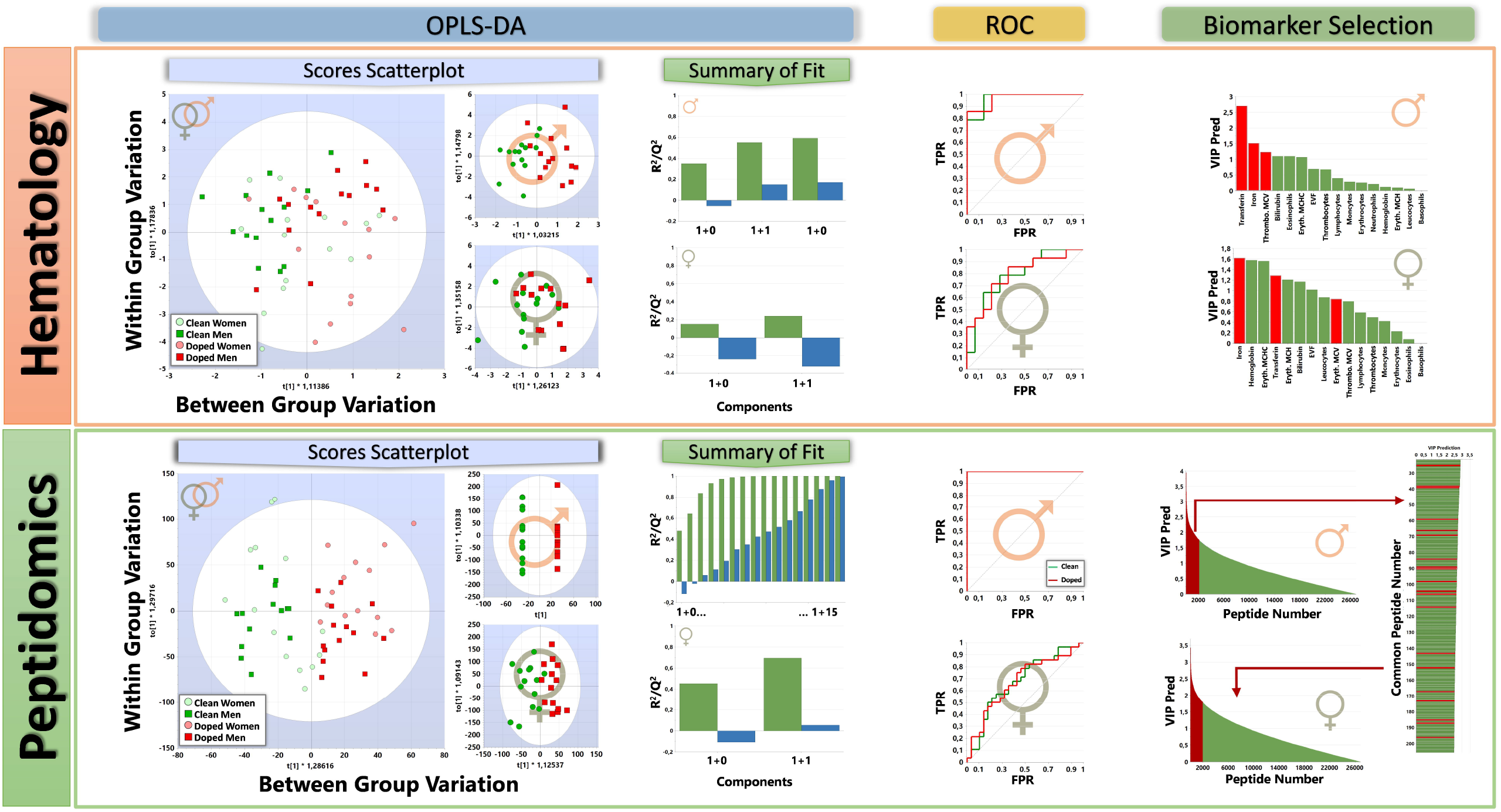
OPLS-DA models

#### Interpretation

Hematological data can generate weak regression models with low predictive power, even when used in combinations.

### Proteomics

Multivariate data analysis (MVA) of quantified and identified peptides from circulating RBC cytosol created significant and cross-validated PCA and OPLS-DA models for regression and prediction of analyzed samples to Clean and Doped groups (Figures 3, Appendix 6). Separation of Clean and Doped samples by OPLS-DA analysis resulted in R^2^ and Q^2^>0.9 for men, and R^2^>0.6 and Q^2^<0.2 for women, displayed in the Summary of Fit plot. Sensitivity and specificity are displayed in the ROC curve, where AUC for Men = 1.00 and Women = 0.68. Selection for prediction by VIP Pred indicate different variables of importance for men and women.

Further results when screening for biomarkers are displayed in Figure 4, where data is divided into three sets; Training (Men), Testing (Women) and Validation (Men and Women). Because the RBC cytosolic fraction (Hb removed) of RBC (RBCc) had different peptidomic profile in men and women, R^2^(cum) = 0.99/Q^2^ = 0.92; P <0.0001, different biomarkers in each sex may be assumed. Models excluding sex-separating peptides were also investigated (Appendix 6).

**FIGURE 4.**
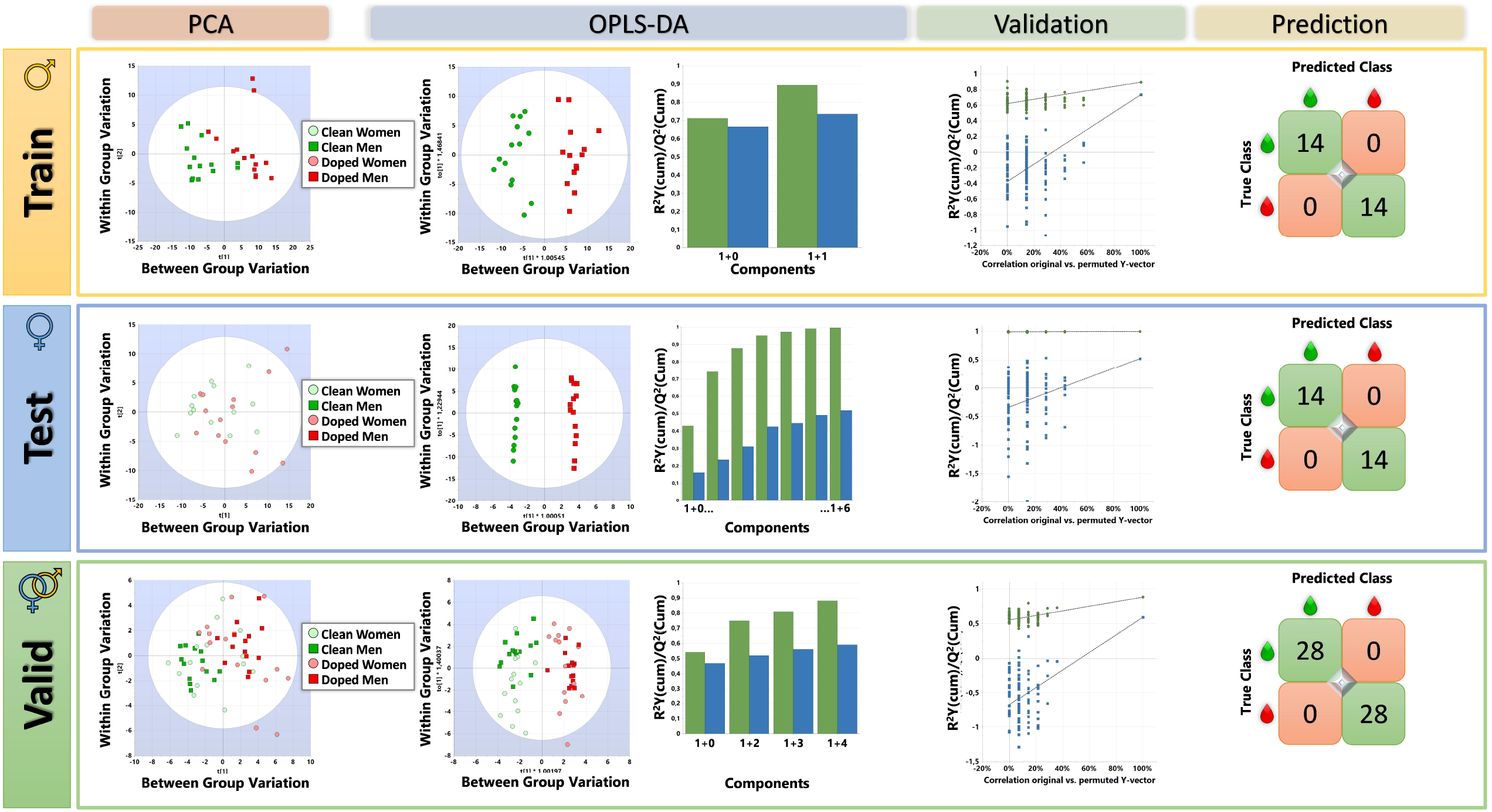
Training/Testing/Validation

#### Training (Men)

the model pared down data to the 200 most important peptides, separating Clean from Doped both in PCA and OPLS-DA analysis, with high R^2^/Q^2^ of 0.90/0.79, cross-validated by 100 permutations and predicting 28/28 samples to the correct class. AUC = 0.96.

#### Testing (Women)

of the model as accomplished by applying the 200 peptides from the Training model on a cohort of women, resulted in R^2^/Q^2^ of 1.00/0.52, cross-validated by 100 permutations and predicting 28/28 samples to the correct class. AUC = 0.92.

#### Validation (Men and Women)

of selected peptides for prediction of Doping used both male and female subjects in combination, starting with the 200 peptides from men and removing peptides until an optimized model was achieved, containing 50 peptides, reaching, R^2^/Q^2^ of 0.88/0.59, cross-validated by 100 permutations and predicting 56/56 samples to the correct class. AUC = 0.97.

Ranking peptides by OPLS-DA loading scores suggests potential candidates for development of diagnostic tools applicable for the detection of autologous blood doping in humans (Figure 5). Merging hematological and peptidomic signature variables, demonstrate a lower loading impact of hematological variables, compared to peptides (Figure 5) in MVA modeling, supporting the previously demonstrated^1^ lack of predictive power using hematology (Figure 1, 2 and 3).

**FIGURE 5.**
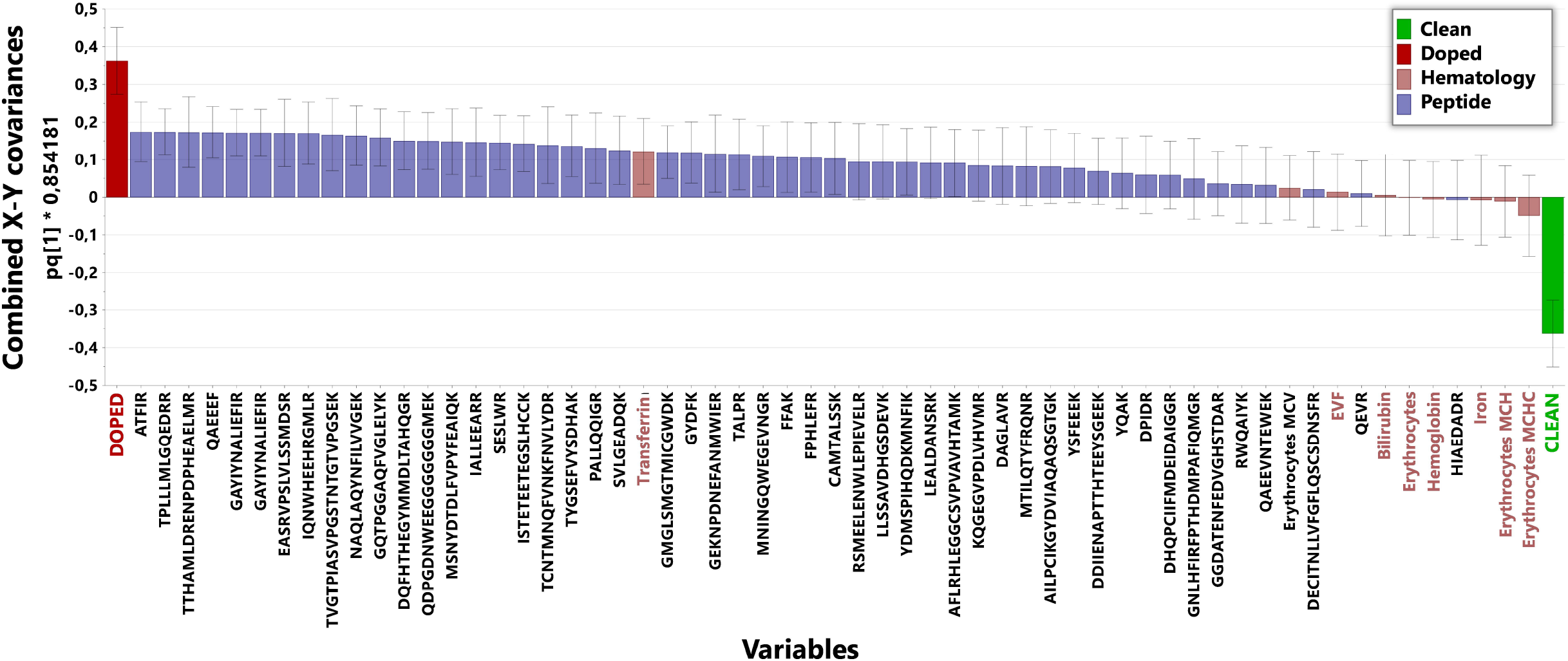
Loading plot

### Confounders

*In vitro* peptides quantified and identified from analysis of the cytosolic fraction of RBCs, harvested from transfusion bags before and after cryo-preservation, created significant and cross-validated OPLS-DA models for the two conditions (Bags, Figure 6). *In vitro* identified peptides did not create significant and cross-validated OPLS-DA models for venous blood samples taken before and after cryo-RBC transfusion (i.e. autologous blood doping, Blood Sample, Figure 6). *In vivo* peptides quantified and identified from analysis of RBCc taken by venous blood sampling in humans before and after cryo-RBC transfusion, created significant and cross-validated OPLS-DA models (Appendix 9/Table 6/Model 19). *In vivo* Doped peptides (top 200 from the OPLS-DA coefficients for Doped) also separated venous blood samples taken from individuals (Elite Athletes) trained at High Altitude, living in Hypoxic Tents and Elite Athlete sampled at sea level (Figure 6, Appendix 9/Table 6). In the Summary plot (Figure 6), samples from Doped do not overlap with other investigated cohorts.

**FIGURE 6.**
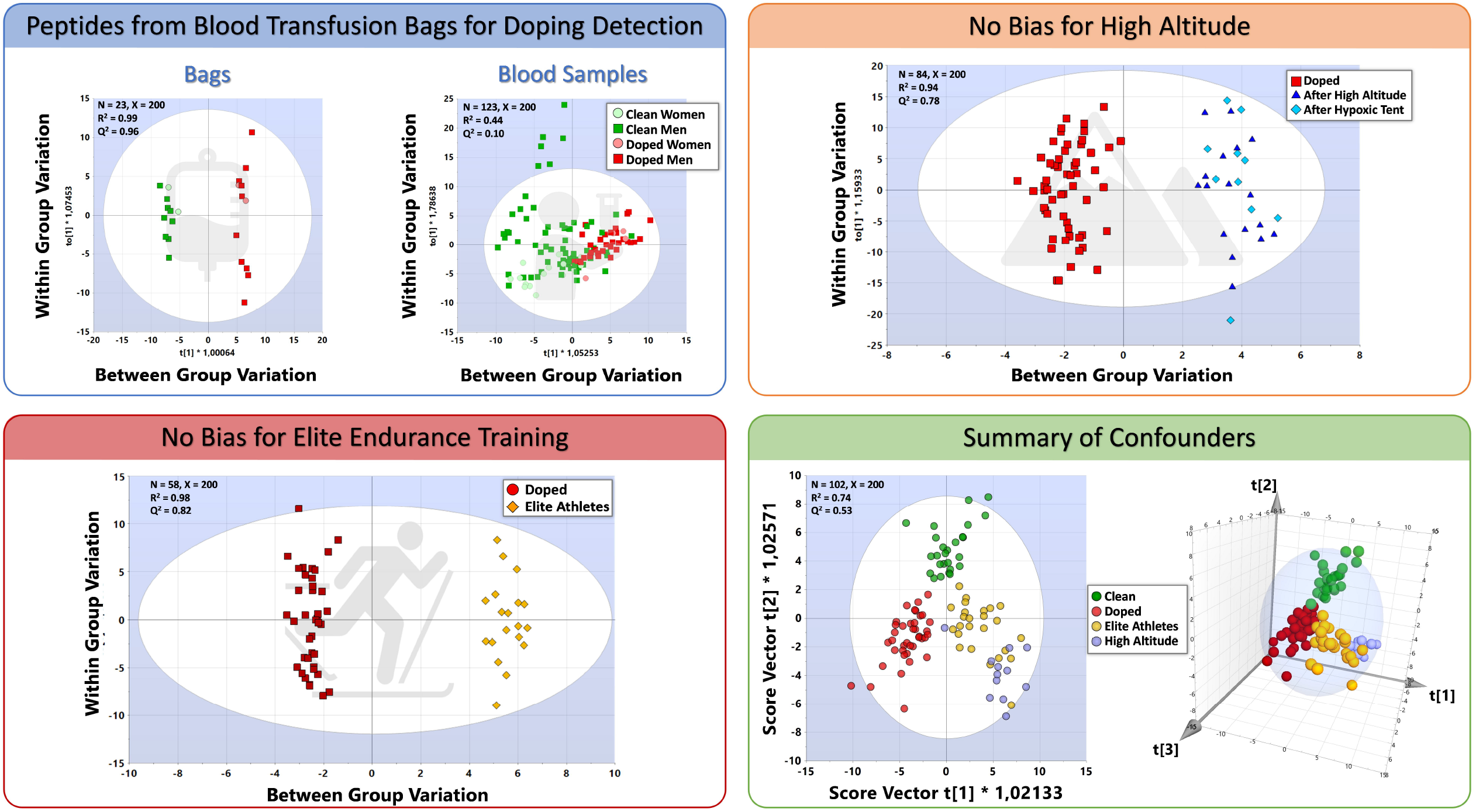
Confounders

#### Interpretation

Proteomic data can generate strong regression models with high predictive power, both for Men and Women, with no impact (bias) from investigated confounders.

## Discussion

Autologous blood doping with cryo-RBC is detected in human subjects by unlabeled, quantitative nano-UPLC-MS/MS. Comparing peptide patterns from RBCc separated Clean from Doped individuals using the multivariate discriminating method OPLS-DA (R^2^>0.90, Q^2^>0.90, p<0.001, X<200). Cross-validated models did not show bias for sex, strenuous physical exercise, hypoxia exposure and high-altitude training in elite athletes. In comparison, hematological variables alone failed to generate significant OPLS-DA models when analyzed in parallel blood samples. Investigations were executed using prospective case-control, cohort and cross-sectional study designs.

### Biomarkers in biological matrixes

Biomarkers for any diagnostics must be reliable, sensitive, specific, and discriminative.^39, 40^ In addition, validation must determine predictability of unknown samples to correct state (i.e. Clean or Doped). For practical implementation in anti-doping efforts to keep sports clean, aspects such as available sample matrices, Doping Control Laboratory (DCL) facilities, transport systems and cost are also important considerations.

Red blood cells are closed systems, unable to synthesis protein and with a life cycle of approximately 120 days in adult humans.^41^ Once set in circulation, RBC undergo an ageing processes, with changing plasma membrane and external structures,^3, 41^ making them susceptible to recognition and subsequent systemic removal by spleen macropahges.^18, 42^ While homologous blood transfusion can be detected via surface antigens,^3^ the only current method to detect autologous blood transfusion is via Hb mass by CO-rebreathing.^43, 44^ The WADA research initiative states that there is no unequivocal detection method for autologous blood doping, as CO is toxic it has no relevance in anti-doping control of healthy athletes who would be reluctant to have a toxic agent imposed on them repeatedly. Autologous blood doping with cryo-RBC is the preferred choice of cheating athletes, as RBC in cold storage (+4°C) has a known clinical safety limit of 42 days after donation, necessitating repeated blood donation, and subsequent decreased performance,^45^ during the preparation phase for competition as the red blood cell population are regenerated to compensate for the loss. Cryo-RBC on the other hand, may be stored for decades.^12^

Regardless of storage procedure, a fraction of all transfused RBCs is damaged and, just as aging RBC, removed from circulation not in 120 days but within 6 h to 48 h.^19^ Consequently, RBC with intact cell surface proteins, and remaining in circulation 48 h after transfusion, may be viewed as circulating time-stamps, containing bioinformation useful in the detection of ABT.^46, 47^ The cryo-induced formation of ice crystals, permanently damaging cytosolic RBC proteins,^21^ serves as a probable, but not the only inducer of potential biomarkers – creating unique detectable peptide fragments after protein trypsinization. An additional advantage of using altered cytosolic biomarkers, compared to hematological variables, is that plasma volume changes will not influence analytical results.

The present series of studies follows a general procedure for biomarker discovery and validation,^39, 40^ including establishing robust baseline values by repeated sample collection at rest in all investigated groups, transports and storage conditions and routine procedures in accredited, clinical laboratories. Current approaches with multiplexing orthogonal biomarkers into one assay increase robustness to confounders and inter-laboratory variations, while reducing analytical costs. The results are reliable, validated for several known possible confounders, have high specificity, sensitivity and discriminating power.^39^

### True positives and Negatives

One unavoidable challenge when investigating ABT is the inclusion of true positives, vital for all statistical analysis and legal actions. As previously directly shown by us^1^, argued by others^23^ and here again demonstrated (Figures 2 and 3) - hematological markers are indirect indicators of ABT, and will always be subject for interpretation (what are limits for normal intra-individual variation?) and significant confounder (exercise, hydration, high altitude, diet, illness etc.) opening for cheating athletes to be legally dismissed from conviction. When investigating hematological intra- and inter-individual variations over time in Clean vs. Doped subjects, neither regression nor prediction models are reliable (Appendix 5). In the context of doping detection, median/range values from elite athletes^22, 31, 48^ (unknown status of doping) are not relevant for individual judgements, as mean/median values from true positively doped subjects^1^ can also stay within 95% CI (Figure 2 & Appendix 5). Also, when using cold stored RBC, detection of ABT by hematological measurements has shown low specificity and sensitivity^31, 49, 50^. Hematological variables changed with investigated confounders, even when more advanced statistical approaches are applied (Figure 1).

The present use of clean elite athletes for comparison is not often possible when validating biomarkers for doping, as athletes cannot serve as subjects. Using cohorts of elite athletes from routine sampling as controls adds uncertainty to any model, as their true state of Clean or Doped cannot be known. Elite athletes participating in this study volunteered to live for extended periods of time in hypoxic tents, before training and competing at high altitude. Knowing they would be tested for blood doping, it is highly unlikely doped athletes are included, also confirmed by the OPLS-DA analysis.

### Window of detection

Theoretically, transfusion of cryo-RBC can be detected for up to 120 days, if the youngest RBCs at time of donation could be identified after transfusion.^19^ Limits of Quantification (LOQ) with the presented method is still not fully investigated, but a few samples (N=14) taken four and five weeks after transfusion (Batch I) are below critical limits (99%) when analyzed by Hotelling’s T^2^ indicating that these samples are no different compared to samples taken one to three weeks after transfusion. Because cryo-induced peptide abundance diminish as RCBs are aged and removed, LOQ should be determined by target analysis methods. Determining LOQ will also be valuable for detection of micro-dosing with cryo-RBC, a practice likely to be used by cheating athletes to avoid detection.

### Alternative interpretations and limitations

An alternative biomarker selection procedure for Clean vs. Doped models could be to exclude the most important (highest OPLS-DA coefficient scores) peptides separating RBCc from Men and Women taken at rest (Clean). This approach may potentially remove peptides important for detecting reinfusion of only one unit cryo-RBC. Creating OPLS-DA models that include 5000 peptides which are most relevant for separating of Men and Women, then selecting the 200 peptides with the highest coefficient score for Doped, creates models for Clean vs. Doped, still biased for sex in the regression models, but without predictive power: Men vs. Women R^2^ = 0.45 and Q^2^ = -1.55, CV-ANOVA p =1.00. Data remaining after removing 5 000 peptides from the model of Men vs. Women, provides an OPLS-DA model for Clean vs. Doped using 200 peptides with R^2^/Q^2^ = 0.64/0.24, CV-ANOVA p < 0.0001. Removing 10 000 peptides gives R^2^/Q^2^ = 0.79/0.38, CV-ANOVA p < 0.001. As discussed, approaching the biomarker selection via a Training set consisting of Men (reinfused with two units RBC) resulted in stronger regression and prediction models (Figure 4). Alternatively, the models for sex investigate cryo-RBC reinfusion of one vs. two units. Data from the present study design cannot distinguish these two options.

Regardless, with the presented list of peptides (Appendix 6 & 9) ABT can be detected in both men and women, reinfused with either one or two units cryo-RBC for at least two weeks after reinfusion.

## Conclusion

- Doping with ABT is directly detected by cryo-induced alterations in cytosolic RBC peptides, with high sensitivity and specificity in both men and women.
- Elite endurance training at low and high altitude did not significantly affect peptides discriminating Clean from Doped samples.
- Presented peptides can be used for direct detection of autologous blood doping using standard equipment available in Doping Control Laboratories.

## Future research

The potential of selected peptides for labeling and direct detection of autologous blood doping by target analysis should be explored, with independent prospective case-control trials and cross-sectional studies, including inter- and intra-laboratory analytical variability. Larger, both-sex cohorts of clean and doped subjects, elite athletes and different ethnic groups need to be sampled for investigation of sensitivity, specificity, false positive and likelihood ratios.

## Data Availability

Data is available in Supplementary files

https://www.dropbox.com/sh/vprl88p6binepmb/AACVJ-mV6jdvqvwDMGDvic30a?dl=0

## Acknowledgements

Outstanding cntributions were given by: The staff at Sunderby Hospital, Luleå for hosting the first series of blood transfusions, under the medical responsibility of Staffan Wikström. The staff at Winternet, Boden for performing all exercise testing and blood sample collection, especially Anna Tano-Nordin and Eva Edholm. The staff at Björknäs Health Center, Boden under guidance of Lillian Falk for organizing the first round of blood donations. The staff at Huddinge Blood Central, where Eva Trollin and Linda Larsson were responsible for blood cryopreservation, and head of unit Hans Gulliksson kindly supported the project. Blood donation, cryopreservation and transfusion, as well as blood sampling, has also been executed by the Center for Stem Cell Apheresis and Management at Huddinge University Hospital, where Emma Watz, Beatrice Asperval Diedrich and Peter Matha managed the study. Ogonna Obudulu for consultation on multivariate statistics. Gunnar Wingsle for technical support with LC-MS/MS, Raik Wagner and Johan Jakobsson for proof-reading and improving the manuscript. All participating subjects for enduring blood loss, exhaustive exercise and numerous trips to the labs.

## Contributors

CM conceived the idea, received funding, developed the methodology and clinical study, performed statistical analysis and was lead writer. IG, MH, PL developed the methodology, prepared samples, executed LC-MS/MS analysis and data cleanup. JL, LG, CH developed the method and executed LC-MS/MS analysis. NK developed the methodology, designed experiments and improved data cleanup. All author participated in writing and editing the final manuscript.

## References

1. Malm CB, Khoo NS, Granlund I, et al. Autologous Doping with Cryopreserved Red Blood Cells - Effects on Physical Performance and Detection by Multivariate Statistics. PLoS One 2016;11(6):e0156157. doi: 10.1371/journal.pone.0156157

2. WADA. Blood doping. https://www.wada-ama.org/en/questions-answers/blood-doping#item-596, 2019.

3. Almizraq R, Tchir JD, Holovati JL, et al. Storage of red blood cells affects membrane composition, microvesiculation, and in vitro quality. Transfusion (Paris) 2013;53(10):2258–67. doi: 10.1111/trf.12080 [published Online First: 2013/01/17]

4. Berglund B, Birgegard G, Wide L, et al. Effects of blood transfusions on some hematological variables in endurance athletes. Med Sci Sports Exerc 1989;21(6):637-42. [published Online First: 1989/12/01]

5. Ekblom B, Goldbarg AN, Gullbring B. Response to exercise after blood loss and reinfusion. J Appl Physiol 1972;33(2):175–80.

6. Lasne F, Martin L, Crepin N, et al. Detection of isoelectric profiles of erythropoietin in urine: differentiation of natural and administered recombinant hormones. Anal Biochem 2002;311(2):119–26. doi: S0003269702004074 [pii] [published Online First: 2002/12/10]

7. Ashenden M, Morkeberg J. Net haemoglobin increase from reinfusion of refrigerated vs. frozen red blood cells after autologous blood transfusions. Vox Sang 2011;101(4):320–6. doi: 10.1111/j.1423-0410.2011.01493.x [published Online First: 2011/05/04]

8. Boning D, Maassen N, Pries A. The Hematocrit Paradox - How Does Blood Doping Really Work? Int J Sports Med 2011;32(4):242–46. doi: 10.1055/s-0030-1255063

9. Schmidt W, Prommer N. Impact of alterations in total hemoglobin mass on VO 2max. Exerc Sport Sci Rev 2010;38(2):68–75. doi: 10.1097/JES.0b013e3181d4957a [published Online First: 2010/03/26]

10. Lundby C, Montero D, Joyner M. Biology of VO2 max: looking under the physiology lamp. Acta Physiol (Oxf) 2017;220(2):218–28. doi: 10.1111/apha.12827 [published Online First: 2016/11/27]

11. Valeri CR, Ragno G, Pivacek LE, et al. An experiment with glycerol-frozen red blood cells stored at -80 degrees C for up to 37 years. Vox Sang 2000;79(3):168–74. doi: 10.1159/000031236 [published Online First: 2000/12/09]

12. MacLare G. Request for the extended storage of frozen red cells beyond 10 years to be considered for inclusion within UK Guidelines. 2016

13. Bizjak DA, Jungen P, Bloch W, et al. Cryopreservation of red blood cells: Effect on rheologic properties and associated metabolic and nitric oxide related parameters. Cryobiology 2018;84:59–68. doi: 10.1016/j.cryobiol.2018.08.001 [published Online First: 2018/08/07]

14. Pallotta V, D’Amici GM, D’Alessandro A, et al. Red blood cell processing for cryopreservation: from fresh blood to deglycerolization. Blood Cells Mol Dis 2012;48(4):226–32. doi: 10.1016/j.bcmd.2012.02.004 [published Online First: 2012/03/20]

15. Walpurgis K, Kohler M, Thomas A, et al. Storage-induced changes of the cytosolic red blood cell proteome analyzed by 2D DIGE and high-resolution/high-accuracy MS. Proteomics 2012;12(21):3263–72. doi: 10.1002/pmic.201200280

16. Holovati JL, Wong KA, Webster JM, et al. The effects of cryopreservation on red blood cell microvesiculation, phosphatidylserine externalization, and CD47 expression. Transfusion (Paris) 2008;48(8):1658–68. doi: 10.1111/j.1537-2995.2008.01735.x [published Online First: 2008/05/17]

17. Hult A, Malm C, Oldenborg PA. Transfusion of cryopreserved human red blood cells into healthy humans is associated with rapid extravascular hemolysis without a proinflammatory cytokine response. Transfusion (Paris) 2013;53(1):28–33. doi: 10.1111/j.1537-2995.2012.03710.x

18. Hult A, Fredrik T, Christer M, et al. Phagocytosis of liquid-stored red blood cells in vitro requires serum and macrophage scavenger receptors. Submitted 2015

19. Mock DM, Matthews NI, Zhu S, et al. Red blood cell (RBC) survival determined in humans using RBCs labeled at multiple biotin densities. Transfusion (Paris) 2011;51(5):1047–57. doi: 10.1111/j.1537-2995.2010.02926.x

20. D’Alessandro A, Gray AD, Szczepiorkowski ZM, et al. Red blood cell metabolic responses to refrigerated storage, rejuvenation, and frozen storage. Transfusion (Paris) 2017;57(4):1019–30. doi: 10.1111/trf.14034 [published Online First: 2017/03/16]

21. Poisson JS, Acker JP, Briard JG, et al. Modulating Intracellular Ice Growth with Cell-Permeating Small-Molecule Ice Recrystallization Inhibitors. Langmuir 2019;35(23):7452–58. doi: 10.1021/acs.langmuir.8b02126 [published Online First: 2018/08/19]

22. Berglund B, Ekblom B, Ekblom E, et al. The Swedish Blood Pass project. Scand J Med Sci Sports 2007;17(3):292–7.

23. Pialoux V, Mounier R, Brugniaux JV. Hemoglobin and hematocrit are not such good candidates to detect autologous blood doping. Int J Hematol 2009;89(5):714–5. doi: 10.1007/s12185-009-0330-5 [published Online First: 2009/05/21]

24. Thevis M, Kohler M, Schanzer W. New drugs and methods of doping and manipulation. Drug discovery today 2008;13(1-2):59–66. doi: S1359-6446(07)00492-8 [pii] 10.1016/j.drudis.2007.11.003 [published Online First: 2008/01/15]

25. Morkeberg JS, Belhage B, Damsgaard R. Changes in blood values in elite cyclist. Int J Sports Med 2009;30(2):130–8. doi: 10.1055/s-2008-1038842 [published Online First: 2008/09/06]

26. Bejder J, Hoffmann MF, Ashenden M, et al. Acute hyperhydration reduces athlete biological passport OFF-hr score. Scand J Med Sci Sports 2015 doi: 10.1111/sms.12438

27. Pottgiesser T, Schumacher YO, Funke H, et al. Gene expression in the detection of autologous blood transfusion in sports--a pilot study. Vox Sang 2009;96(4):333–6. doi: VOX1169 [pii] 10.1111/j.1423-0410.2009.01169.x [published Online First: 2009/03/04]

28. Gore CJ, Parisotto R, Ashenden MJ, et al. Second-generation blood tests to detect erythropoietin abuse by athletes. Haematologica 2003;88(3):333–44. [published Online First: 2003/03/26]

29. Monfort N, Ventura R, Latorre A, et al. Urinary di-(2-ethylhexyl)phthalate metabolites in athletes as screening measure for illicit blood doping: a comparison study with patients receiving blood transfusion. Transfusion (Paris) 2009;50(1):145–9. doi: TRF2352 [pii] 10.1111/j.1537-2995.2009.02352.x [published Online First: 2009/08/22]

30. Pottgiesser T, Umhau M, Ahlgrim C, et al. Hb mass measurement suitable to screen for illicit autologous blood transfusions. Med Sci Sports Exerc 2007;39(10):1748–56. doi: 10.1249/mss.0b013e318123e8a6 00005768-200710000-00011 [pii] [published Online First: 2007/10/03]

31. Morkeberg J, Sharpe K, Belhage B, et al. Detecting autologous blood transfusions: a comparison of three passport approaches and four blood markers. Scand J Med Sci Sports 2011;21(2):235–43. doi: 10.1111/j.1600-0838.2009.01033.x [published Online First: 2009/11/12]

32. Thevis M, Kuuranne T, Walpurgis K, et al. Annual banned-substance review: analytical approaches in human sports drug testing. Drug Test Anal 2016;8(1):7–29. doi: 10.1002/dta.1928

33. Fitch KD. Blood doping at the Olympic Games. J Sport Med Phys Fit 2017;57(11):1526–32. doi: 10.23736/S0022-4707.17.06948-1

34. Al-Thani AM, Voss SC, Al-Menhali AS, et al. Whole Blood Storage in CPDA1 Blood Bags Alters Erythrocyte Membrane Proteome. Oxid Med Cell Longev 2018;2018:6375379. doi: 10.1155/2018/6375379 [published Online First: 2018/12/12]

35. Bylesjo M, Eriksson D, Sjodin A, et al. Orthogonal projections to latent structures as a strategy for microarray data normalization. BMC Bioinformatics 2007;8:207. doi: 1471-2105-8-207 [pii] 10.1186/1471-2105-8-207 [published Online First: 2007/06/20]

36. Pohjanen E, Thysell E, Jonsson P, et al. A multivariate screening strategy for investigating metabolic effects of strenuous physical exercise in human serum. J Proteome Res 2007;6(6):2113–20. doi: 10.1021/pr070007g [published Online First: 2007/04/13]

37. Wold S, Esbensen K, Geladi P. Principal Component Analysis. Chemometrics and Intelligent Laboratory Systems 1987;2:37–52.

38. Trygg J, Holmes E, Lundstedt T. Chemometrics in metabonomics. J Proteome Res 2007;6(2):469–79. doi: 10.1021/pr060594q [published Online First: 2007/02/03]

39. Aikin R, Baume N, Equey T, et al. Biomarkers of doping: uses, discovery and validation. Bioanalysis 2020 doi: 10.4155/bio-2020-0035 [published Online First: 2020/06/02]

40. Brun V, Couté Y, editors. Proteomics for Biomarker Discovery: Methods and Protocols. Université Grenoble Alpes, CEA, Inserm, BGE U1038GrenobleFrance: Human Press, 2019.

41. Bosman GJ. Survival of red blood cells after transfusion: processes and consequences. Front Physiol 2013;4:376. doi: 10.3389/fphys.2013.00376

42. Hult A. Towards a detailed understanding of the red blood cell storage lesion: and its consequences for in vivo survival following transfusion. Umeå Universiy, 2015.

43. Schmidt W, Prommer N. The optimised CO-rebreathing method: a new tool to determine total haemoglobin mass routinely. Eur J Appl Physiol 2005;95(5-6):486–95. doi: 10.1007/s00421-005-0050-3 [published Online First: 2005/10/14]

44. Siebenmann C, Keiser S, Robach P, et al. CORP: The assessment of total hemoglobin mass by carbon monoxide rebreathing. J Appl Physiol 2017;123(3):645–54. doi: 10.1152/japplphysiol.00185.2017 [published Online First: 2017/07/01]

45. Celsing F, Nystrom J, Pihlstedt P, et al. Effect of long-term anemia and retransfusion on central circulation during exercise. J Appl Physiol (1985) 1986;61(4):1358–62.

46. Karsten E, Breen E, Herbert BR. Red blood cells are dynamic reservoirs of cytokines. Sci Rep 2018;8(1):3101. doi: 10.1038/s41598-018-21387-w

47. Carvalho AS, Rodriguez MS, Matthiesen R. Red Blood Cells in Clinical Proteomics. Methods Mol Biol 2017;1619:173–81. doi: 10.1007/978-1-4939-7057-5_13

48. Malcovati L, Pascutto C, Cazzola M. Hematologic passport for athletes competing in endurance sports: a feasibility study. Haematologica 2003;88(5):570–81.

49. Pottgiesser T, Echteler T, Sottas PE, et al. Hemoglobin mass and biological passport for the detection of autologous blood doping. Med Sci Sports Exerc 2012;44(5):835–43. doi: 10.1249/MSS.0b013e31823bcfb6

50. Sallet P, Brunet-Guedj E, Mornex R, et al. Study of a new indirect method based on absolute norms of variation to detect autologous blood transfusion. Int J Hematol 2008;88(4):362–8. doi: 10.1007/s12185-008-0182-4 [published Online First: 2008/10/24]

